# A Systematic Review and Meta-Analysis of Studies Examining the Relationship Between Reported Racism and Health and Wellbeing for Children and Adolescents

**DOI:** 10.1101/2020.12.02.20243022

**Authors:** Naomi Priest, Kate Doery, Mandy Truong, Shuaijun Guo, Brigid Trenerry, Saffron Karlsen, Yvonne Kelly, Yin Paradies

**Affiliations:** Australian National University, Centre for Social Research and Methods, Canberra, Australia; Murdoch Children’s Research Centre, Population Health, Melbourne, Australia; Monash University, School of Nursing and Midwifery, Melbourne, Australia; University of Melbourne, Department of Pediatrics, Melbourne, Australia; Singapore University of Technology and Design, Lee Kuan Yew Centre for Innovative Cities, Singapore; University of Bristol, School of Sociology, Politics and International Studies, Bristol, United Kingdom; University College London, Department of Epidemiology and Public Health, London, United Kingdom; Deakin University, School of Humanities and Social Science, Melbourne, Australia

**Keywords:** Protocol, Systematic-Review, Meta-analysis, Racism, Child Health

## Abstract

**Introduction:** There is a growing body of research showing associations between experiences of racism and poor health and wellbeing outcomes for children and adolescents. The aim of this review protocol is to update the first systematic review conducted by Priest et al. 2013, including a meta-analysis of findings. Based on previous empirical data, it is anticipated that child and adolescent health will be negatively impacted by racism. This review will provide updated evidence of effect sizes across outcomes and identify moderators and mediators of relationships.

**Methods and analysis:** This systematic review and meta-analysis will include studies that explore associations between experiences of racism and racial dissemination with health outcomes of children and adolescents aged 0- 24 years of age from any racial/ethnic/cultural group. Outcome measures include general health and wellbeing, physical health, mental health, healthcare utilisation and health behaviours. Exposure measures include self-reported and proxy reported personal experiences of racism and reported experiences of vicarious racism. The authors will conduct a comprehensive search of studies from the earliest time available to September 2020. All relevant studies will be screened with data extraction, quality appraisal and publication bias conducted independently by at least two authors.

**Ethics and Dissemination:** This review will provide evidence for future research within the field and help to support policy and practice development. Results from this systematic review and meta-analysis will be widely disseminated to both academic and non-academic audiences.

Ethics approval is not required as this is a review of existing empirical findings.

**Article Summary:**

- This is an updated systematic review which aims to update the findings from the first international review conducted by Priest et al. 2013. However, it is the first meta-analysis to be conducted exploring the relationship between racism and health in child and adolescent from all ethnic/racial/cultural backgrounds.
- This systematic review will show the health effects of racial discrimination on child and adolescent health, the key pathways by which racial discrimination influences these outcomes and identify potential moderators and mediators.
- Findings from this systematic review and meta-analysis will be used to provide recommendations for future research and inform the development of effective evidence-based strategies for addressing racism and ameliorating its harmful effects.
- This systematic review has a bias towards papers published in English as this review will only search studies published in English, meaning that studies not-published in English will not be included in this review. By doing so this review may not include all findings of all relevant studies.

## INTRODUCTION

Racism and racial discrimination are widely recognised as critical determinants of health and health inequities for children and adolescents across populations and contexts^1-3^. Racism is a system of oppression that categorises and stratifies social groups into ‘races’, devalues and disadvantages those considered inferior and differentially allocates to them valued societal resources and opportunities^4 5^. Racism is expressed across multiple levels, including systemic and inter- and intra-personal levels, and operates in many forms including vicarious racism, whereby individuals experience racism on a secondary level, witnessing or being informed of family, friends and strangers experiencing racism^6 7^. Racial discrimination is the behavioural expression of racism and also manifests at systemic and individual levels. Currently the world is currently experiencing two global pandemics, COVID- 19 and racism, with many people experiencing heightened levels of discrimination which is having negative impacts on people’ health and wellbeing^8^.

Research on racism and health has predominantly focused on interpersonal experiences, with considerable evidence documenting negative health effects across multiple outcomes^5 9 10^. However, most of this evidence remains focusing on adults, with far less research conducted among children and adolescents.

Priest et al.^1^ conducted the first international systematic review of epidemiological studies on reported racial discrimination and the health and wellbeing of children and adolescents, including 121 studies. However since this report was published in 2013 the contribution of racism as a social determinant of health and wellbeing among children and adolescents has received growing attention^3^. There is increasing evidence of the impact of racism on pathophysiological processes (e.g. allostatic load and stress neurobiology) and biological markers (e.g. C-reactive protein and cortisol)^11^ as well as on sleep^12-14^ among children and adolescents.

A recent review of vicarious racism and child health found 30 studies published up to May 2016 compared with 10 studies in the previous 2013 review (with studies searched up to November 2011)^6^. This represents a three-fold increase in studies examining vicarious racism and child health in approximately four and a half years. Additionally, our original review found that two-thirds of the included studies were published between 2005- 2012^1^. In light of the burgeoning research in the field, there is a need to review and reflect on the current evidence to inform future scholarship in this area.

This present systematic review and meta-analysis aims to update findings from the 2013 review conducted by Priest et al.^1^ An updated systematic review is necessary to include new data, new methods, and updated analysis to a similar research question.^15^ In this instance an updated systematic review is necessary due to changing social policy and demographic contexts and new health priorities globally, as well as an increase in the number of recent publications in this area, including in different country and population contexts. The first systematic review identified that there were a limited number of longitudinal studies that have explored the health effects of racism on children and a need to expand research in this area, with a focus needed on the complex pathways to which child and youth health is impacted by experiences of racial discrimination^1^. Priest et al. called for an increase in high-quality longitudinal research utilising robust multidimensional measures of racial discrimination^1^. As highlighted since this review was published in 2013, there has been a large increase in the amount of research being conducted in this field.

This present systematic review and meta-analysis will use the previous review as a guide, building upon it and utilising an updated inclusion and exclusion strategy as well as expanding it to include a meta-analysis. As indicated by Garner et al.^15^ an updated systematic review can have an updated inclusion criteria whilst answering a similar question.

The broad aims of this present systematic review and meta-analysis are:

- To further understanding of the health effects of racial discrimination on child and adolescent health, the key pathways by which racial discrimination influences these outcomes and identify potential moderators and mediators.
- To provide key recommendations for future research and inform the development of effective evidence-based strategies for addressing racism and ameliorating its harmful effects.

## METHODS AND ANALYSIS

This systemic review and meta-analysis will follow the guidelines of the Preferred Reporting Items for Systematic Reviews and Meta-Analysis (PRISMA)^16^ with the PRISMA Protocols (PRISMA-P)^17^ checklist followed for the writing of this protocol and is included as appendix 1.

Progress on this systematic review and meta-analysis will be updated on the International Prospective Register of Systematic Reviews (PROSPERO) to maintain transparency.

### Inclusion criteria

Studies using quantitative methods including cross-sectional; prospective and retrospective cohort; and case– control designs will be included. Peer-reviewed journal articles (published or available as pre-prints), and dissertations/theses will be included. We will also include grey literature including published reports. Studies that do not report empirical associations between racism and child and adolescent health will not be included.

### Participants

Participants will include children and adolescents aged up to 24 years from any racial/ethnic/cultural groups. The age range of participants has been updated since the previous review (which included participants up to 18 years) due to a shift in the definition of the age of adolescence.^18^

### Exposure

This review will include studies which focus on reported childhood experiences of racism as the exposure, including: self-reported racism, proxy reports from child’s experiences or racism as reported by parents or carers including experiences of vicarious racism (for example, witnessing racism experienced by family or friends). There will be no restrictions placed on the timeframe of exposure to racism prior to the measurement. Retrospective adult population studies that report on childhood experiences of racism will be noted, but will not be included in our analysis.

### Outcome measures

Studies will be considered if they measure health outcomes in children and adolescents. Health and wellbeing outcomes include measures of ill-health and illness as well as positive health outcomes across physical, mental and behavioural domains. As guided by previous reviews and research^1 9 12 19-24^, the following health and wellbeing outcomes will be included:

1. Pregnancy and birth outcomes (e.g. premature birth, low birth weight)
2. General health and wellbeing
3. Physical health (infectious disease and chronic conditions and markers e.g. body mass index, waist hip ratio, blood pressure, metabolic and cardiovascular disease, overweight, obesity)
4. Mental health (e.g. social and emotional difficulties, psychological adjustment and distress, self-esteem, mental illness, suicide risk, sleep difficulties, psychosis)
5. Wellbeing, life satisfaction, quality of life, resilience
6. Positive mental health (e.g. self-esteem, self-worth and resilience)
7. Learning and developmental difficulties (e.g. developmental delay, concerns about learning, externalising behaviours such as violence etc., poor attachment etc.)
8. Health/risk behaviours (e.g. alcohol, tobacco, substance use)
9. Health care utilisation, healthcare costs, satisfaction with child health care system (use of screening tests, maternal child health care, access to health care and treatment, adherence to treatment)
10. Biological markers (e.g. inflammation and cardiometabolic markers)

### Exclusion criteria

Studies reporting the effects of reported racism on other outcomes (e.g. education, employment) will not be included. Studies where specific effects of racism cannot be isolated from broader measure of discrimination will be noted but will not be included in the meta-analysis. Only studies published in English will be included.

Qualitative studies or studies only reporting the prevalence of racism without identifying associations with health and wellbeing outcomes will not be included.

### Data extraction and management

#### Search Strategy

The search strategy will be conducted in English and include studies from the earliest time available to September 2020. The search strategy will not be restricted to papers only published since the completion of the previous search strategy in 2011 as databases regularly back index studies and therefore some studies may have been missed by the original review. The search will be checked against the original search results to ensure that all studies that have been back indexed are also included.

The search will be conducted in the Ovid Medline, Ovid PsycInfo, PubMed, ERIC and ProQuest (for dissertation/theses) databases. Reference lists will also be hand-searched for relevant studies. The authors will also search google scholar and key websites as well as contacting experts within the field to make sure all relevant studies including grey literature are included.

The search will be performed using a string template combining search terms relevant to our study population, exposure and outcomes. The search strategy template has been developed in consultation with medical library staff utilising the previous search strategy by Priest et al.^1^ as a template. The search template to be used for Medline is included as appendix 2, which will be updated accordingly for each database.

#### Selection of studies

One member of the review team will conduct the initial search in the selected databases with the search results to be imported into Endnote X9^25^, with duplicates and papers not in English to be deleted. All titles and abstracts of studies identified in the search will be independently screened for eligibility for inclusion by members of the review team using Covidence. Full text studies will be assessed for final inclusion. Any discrepancies between members of the review team will be resolved by having a third member of the review team adjudicate the decision. Rationale for exclusion of studies will be noted throughout the screening process with a PRISMA flowchart^16^ being used to show the full selection process of studies.

Further, studies with inappropriate and/or insufficient data to allow meta-analysis will be documented, but excluded from the quantitative analysis.

#### Data Extraction

Once the full text studies have been identified, members of the review team will extract the data using Airtable^26^. An independent member of the review team will compare and check for inconsistencies with discrepancies resolved through discussion. Data from some studies may appear in multiple publications. If publications include unique combinations of exposure and outcome variables they will be extracted as distinct data sets; meaning that one study may be included in the meta-analysis multiple times as different data-sets due to its use of multiple measures of health or racism.

Specifically, this review will examine the key characteristics of studies of reported racism and health and wellbeing among children and adolescents including:

- where and when studies have been conducted, the racial/cultural/ethnic background, age and gender of study populations, study designs, sample sizes, and data sources used;
- how reported racism is defined, how this exposure is measured in terms of method of administration, content and time frames of exposure, targets and perpetrators of racism, reactions/responses to racism and settings in which racism is experienced; and
- the prevalence of reported racism and direction/magnitude/effect sizes of associations found between reported racism and health across health outcomes, study and exposure characteristics and identify effect moderator and mediators of these associations.

Data to be extracted will include: authors; year of publication; study aims; study design (including sampling methods); definition of racism exposure measure (s) (including tools/instruments and psychometric properties when applicable; method of administration including informant(s); content and time frames of exposure, targets and perpetrators of racism, reactions/responses to racism and settings in which racism is experienced); health outcome measures; measure of racial/cultural/ethnic background; study location (country/region); place of residence (urban/rural), sample size; participant demographics (age, racial/cultural backgrounds, gender, religion, education, socio-economic status, migration status); study findings; prevalence of self-reported racism including exposure characteristics; nature of associations between self-reported racism and health and wellbeing outcomes (including subgroup analysis when reported (mean, SD, effect size)); reactions and responses to racism; confounders, effect moderator and mediators of these associations; study recommendations; and study quality/critical appraisal.

Effect sizes such as coefficients and p-values for each health outcome will be extracted. Both unadjusted and adjusted effect sizes will be extracted when available, and covariates included in models recorded. Only effect sizes for children and adolescents will be extracted. Where an overall effect size is reported across a range of ages, we will extract subgroup effect sizes when reported.

### Assessment of study quality and bias

Studies included in the review will be critically appraised to determine the validity of the study’s findings from the known literature and to provide readers with the ability to make an informed decision on the quality of these findings. The review team will use a variety of Critical Appraisal Tools, utilising tools that are most relevant to the methods used in the study. We will utilise the Systematic Review Toolbox to assist with finding the most appropriate tools.^27^ These tools will include the Newcastle- Ottawa Scale^28^ and the Joanna Briggs critical appraisal tools appropriate to relevant study designs. Any discrepancies between quality ratings will be resolved through discussion between members of the review team. The quality ratings will be used as a guide for data analysis and meta-analysis.

The review team will also assess the publication bias of the included studies. The review team will use a variety of measures to assess the publication bias of the included studies, these will include using the Risk Of Bias In Non-randomized Studies - of Exposures (ROBINS-E) Tool^29^, an assessment tool under development which assesses seven different domains of bias in exposure studies,^30^ as included as Appendix 3.

### Analysis

Data that meet all inclusion criteria will first be summarised descriptively and then analysed statistically. Data analysis will be conducted using Airtable^26^.

A descriptive summary with data tables will be produced to summarise the literature. Study characteristics will be presented in summary tables across key variables (including their specific design, study details, sample size, age and racial or ethnic background of participants, exposure and outcome measures included).

Although meta-analysis is planned this will only become apparent when extracted data are reviewed for feasibility. If data is available we will conduct analyses of associations between racism and health for different health outcome measures, and at different time points. If possible, we will use random-effects models to aggregate effect sizes. Subgroup analyses will be conducted for age, gender and ethnicity if possible. To assess the heterogeneity of studies, we will use the Q-statistic test and the I^2^ statistic. If the test for heterogeneity denoted as I^2^ (if I^2^≤25%), studies will be considered homogeneous.

## DISCUSSION

As this is an updated systematic review and meta-analysis, we expect that whilst there has been a significant amount of recent research conducted in this space, we do not anticipate our findings to be vastly different from our original review. This review and meta-analysis will incorporate studies with participants from all ethnic/racial/cultural backgrounds, and studies will not be limited to any one country or geographic area, and in doing so we anticipate to show that this is a problem facing not just one specific population but children globally. That is, we expect the review to show that racism and racial discrimination negatively impact on multiple health outcomes in children and adolescents from different ethnic/racial/cultural backgrounds and across contexts. We expect an increase of research in outcomes not considered in the original review, including sleep and inflammatory and immune biomarkers, as well as markers of epigenetic risk and cellular aging and of endocrine and hormonal function. Increased attention on younger age groups, vicarious as well as direct exposure, longitudinal associations, and populations and settings outside of the United States of America is also anticipated.

The world is currently facing two global pandemics, with high levels of racial discrimination being experienced globally throughout the COVID-19 pandemic^8^, and racism has been drawing increased media attention. Exploring the health effects of racism and discrimination is paramount. Due to the expectant increase in research surrounding this topic, a key contribution of the current study is to conduct a meta-analysis, which was not able to be conducted before. We expect that through this meta-analysis we will be able to show rigorous and robust evidence showing the relationship between experiences of racism and health and wellbeing outcomes for children and adolescents. As this is the first meta-analysis of these studies, it will provide an evidence base for future research exploring the effect of racism and child health, as well as for policy development and service delivery.

## Data Availability

This is a protocol paper for a systematic review and Meta-analysis.

## ETHICS AND DISSEMINATION

Ethics approval is not required as this is a review of existing empirical findings and no primary data will be collected throughout the research.

The results from this review will be disseminated in peer-review publications, conference presentations as well as communicated more broadly through factsheets and summaries disseminated through academic institution press release and policy and practice partners.

Progress on this systematic review and meta-analysis will be updated on the International Prospective Register of Systematic Reviews (PROSPERO) (PROSPERO ID 184055) to maintain transparency.

## AUTHOR CONTRIBUTIONS

NP conceptualised the review and contributed to all aspects of the protocol. KD drafted the protocol, developed the search strategy with medical librarians. MT, JG, BT, SK, YK and YP reviewed the protocol draft.

## REGISTRATION

The systematic review will be registered with PROSPERO, ID number 184055.

## Appendix 1 PRISMA-P (Preferred Reporting Items for Systematic review and Meta-Analysis Protocols) 2015 checklist: recommended items to address in a systematic review protocol*

| Section and topic | Item No | Checklist item |
| --- | --- | --- |
| **ADMINISTRATIVE INFORMATION** |  |  |
| Title: |  |  |
| Identification | 1a | Identify the report as a protocol of a systematic review |
| Update | 1b | If the protocol is for an update of a previous systematic review, identify as such |
| Registration | 2 | If registered, provide the name of the registry (such as PROSPERO) and registration number |
| Authors: |  |  |
| Contact | 3a | Provide name, institutional affiliation, e-mail address of all protocol authors; provide physical mailing address of corresponding author |
| Contributions | 3b | Describe contributions of protocol authors and identify the guarantor of the review |
| Amendments | 4 | If the protocol represents an amendment of a previously completed or published protocol, identify as such and list changes; otherwise, state plan for documenting important protocol amendments |
| Support: |  |  |
| Sources | 5a | Indicate sources of financial or other support for the review |
| Sponsor | 5b | Provide name for the review funder and/or sponsor |
| Role of sponsor or funder | 5c | Describe roles of funder(s), sponsor(s), and/or institution(s), if any, in developing the protocol |
| **INTRODUCTION** |  |  |
| Rationale | 6 | Describe the rationale for the review in the context of what is already known |
| Objectives | 7 | Provide an explicit statement of the question(s) the review will address with reference to participants, interventions, comparators, and outcomes (PICO) |
| **METHODS** |  |  |
| Eligibility criteria | 8 | Specify the study characteristics (such as PICO, study design, setting, time frame) and report characteristics (such as years considered, language, publication status) to be used as criteria for eligibility for the review |
| Information sources | 9 | Describe all intended information sources (such as electronic databases, contact with study authors, trial registers or other grey literature sources) with planned dates of coverage |
| Search strategy | 10 | Present draft of search strategy to be used for at least one electronic database, including planned limits, such that it could be repeated |
| Study records: |  |  |
| Data management | 11a | Describe the mechanism(s) that will be used to manage records and data throughout the review |
| Selection process | 11b | State the process that will be used for selecting studies (such as two independent reviewers) through each phase of the review (that is, screening, eligibility and inclusion in meta-analysis) |
| Data collection process | 11c | Describe planned method of extracting data from reports (such as piloting forms, done independently, in duplicate), any processes for obtaining and confirming data from investigators |
| Data items | 12 | List and define all variables for which data will be sought (such as PICO items, funding sources), any pre-planned data assumptions and simplifications |
| Outcomes and prioritization | 13 | List and define all outcomes for which data will be sought, including prioritization of main and additional outcomes, with rationale |
| Risk of bias in individual studies | 14 | Describe anticipated methods for assessing risk of bias of individual studies, including whether this will be done at the outcome or study level, or both; state how this information will be used in data synthesis |
| Data synthesis | 15a | Describe criteria under which study data will be quantitatively synthesised |
| | 15b | If data are appropriate for quantitative synthesis, describe planned summary measures, methods of handling data and methods of combining data from studies, including any planned exploration of consistency (such as $I^2$, Kendall's $\tau$) |
|  | 15c | Describe any proposed additional analyses (such as sensitivity or subgroup analyses, meta-regression) |
|  | 15d | If quantitative synthesis is not appropriate, describe the type of summary planned |
| Meta-bias(es) | 16 | Specify any planned assessment of meta-bias(es) (such as publication bias across studies, selective reporting within studies) |
| Confidence in cumulative evidence | 17 | Describe how the strength of the body of evidence will be assessed (such as GRADE) |

**\* It is strongly recommended that this checklist be read in conjunction with the PRISMA-P Explanation and Elaboration (cite when available) for important clarification on the items. Amendments to a review protocol should be tracked and dated. The copyright for PRISMA-P (including checklist) is held by the PRISMA-P Group and is distributed under a Creative Commons Attribution Licence 4.0.**

*From: Shamseer L, Moher D, Clarke M, Ghersi D, Liberati A, Petticrew M, Shekelle P, Stewart L, PRISMA-P Group. Preferred reporting items for systematic review and meta-analysis protocols (PRISMA-P) 2015: elaboration and explanation. BMJ. 2015 Jan 2;349(jan02 1):g7647*.

## Appendix 2 Search Strategy Search date: 18/7/2020

**Database(s): Ovid MEDLINE(R) ALL 1946 to July 16, 2020**

Search Strategy:

1. Prejudice/ or Racism/
2. (racism or racial-discriminat* or racial-prejudice or racist-event* or racist-episode* or racial-stereotype* or race-related-stress).tw,kf.
3. ((discriminat* or bias* or prejudic* or hostil* or harass* or bully* or cyberbull* or cyber-bull* or (unfair* adj1 treat*) or oppress*) adj3 (race or racial* or ethnic* or cultur* or religio* or migrant* or refugee* or asylum)).tw,kf.
4. (newborn* or new-born* or baby or babies or neonat* or neo-nat* or infan* or toddler* or pre-schooler* or preschooler* or kinder or kinders or kindergarten* or boy or boys or girl or girls or child or children or childhood or pediatric* or paediatric* or adolescen* or youth or youths or teen or teens or teenage* or school-age* or schoolage* or school-child* or schoolchild* or school-girl* or schoolgirl* or school-boy* or schoolboy* or young-person* or young-people).af.
5. Child Welfare/ or pediatric obesity/et, ep, pc
6. (Prejudice/ or *Racism/ or 2 or 3) and 5
7. obesity/et, ep, pc or body mass index/ or overweight/pc
8. Waist-Hip Ratio/
9. Blood Pressure/ or Biomarkers/
10. Hypertension/et, ep, pc
11. exp Cardiovascular Diseases/et, ep, pc
12. depression/et, ep, pc or anxiety/et, ep, pc
13. Mental Health/
14. Stress, Psychological/et, ep, pc
15. Sleep/
16. exp Sleep Wake Disorders/et, ep, pc
17. “Quality of Life”/
18. Resilience, Psychological/ or exp adaptation, psychological/
19. exp substance-related disorders/et, ep, pc or alcohol-related disorders/et, ep, pc
20. smoking/et, ep or exp tobacco smoking/et, ep, pc
21. Mental Disorders/et, ep, pc
22. Self Concept/
23. personal satisfaction/
24. exp suicide/et, ep, pc
25. conduct disorder/et, ep, pc or aggression/et, ep, pc
26. pregnancy outcome/
27. (health-care or healthcare or health-service* or clinic? or ill-health or wellbeing or well-being or disease* or illness* or bmi or body-mass-index or anthropometric* or WHR or waist-hip-ratio or hypertension or blood-pressure or cardiometabolic or cardio-metabolic or biomarker* or obese or obesity or overweight or depress* or anxiety or anxious* or mental-health or mental-disorder* or stress or distress* or suicid* or sleep or psychosis or tobacco or smoke* or smoking or drug? or alcohol* or substance-use or substance-related-disorder* or resilien* or self-esteem or self-worth or self-concept or quality-of-life or life-satisfaction or personal-satisfaction or conduct-disorder* or aggression).tw,kf.
28. ((social or behavio* or emotion* or developmental* or psychological* or learning*) adj3 (difficul* or problem* or delay* or adjust*)).tw,kf.
29. (((pregnancy or birth or gestation*) and (outcome* or preterm or pre-term or premature or small-for-gestational-age)) or low-birthweight or low-birth-weight).tw,kf.
30. 7 or 8 or 9 or 10 or 11 or 12 or 13 or 14 or 15 or 16 or 17 or 18 or 19 or 20 or 21 or 22 or 23 or 24 or 25 or 26
31. *obesity/et, ep, pc or *body mass index/ or *overweight/pc or *Waist-Hip Ratio/ or (*Blood Pressure/ or *Biomarkers/) or *Hypertension/et, ep, pc or exp *Cardiovascular Diseases/et, ep, pc or (*depression/et, ep, pc or *anxiety/et, ep, pc) or *Mental Health/ or *Stress, Psychological/et, ep, pc or *Sleep/ or exp *Sleep Wake Disorders/et, ep, pc or *”Quality of Life”/ or (*Resilience, Psychological/ or exp *adaptation, psychological/) or (exp *substance-related disorders/et, ep, pc or *alcohol-related disorders/et, ep, pc) or (*smoking/et, ep or exp *tobacco smoking/et, ep, pc) or *Mental Disorders/et, ep, pc or *Self Concept/ or *personal satisfaction/ or exp *suicide/et, ep, pc or (*conduct disorder/et, ep, pc or *aggression/et, ep, pc) or *pregnancy outcome/
32. (Prejudice/ or *Racism/ or 2 or 3) and (27 or 28 or 29 or 31) and 4
33. 33 6 or 32
34. 34 limit 33 to (comment or editorial or letter)
35. **35 33 not 34**

## Appendix 3: The ROBINS-E tool (Risk Of Bias In Non-randomized Studies - of Exposures)

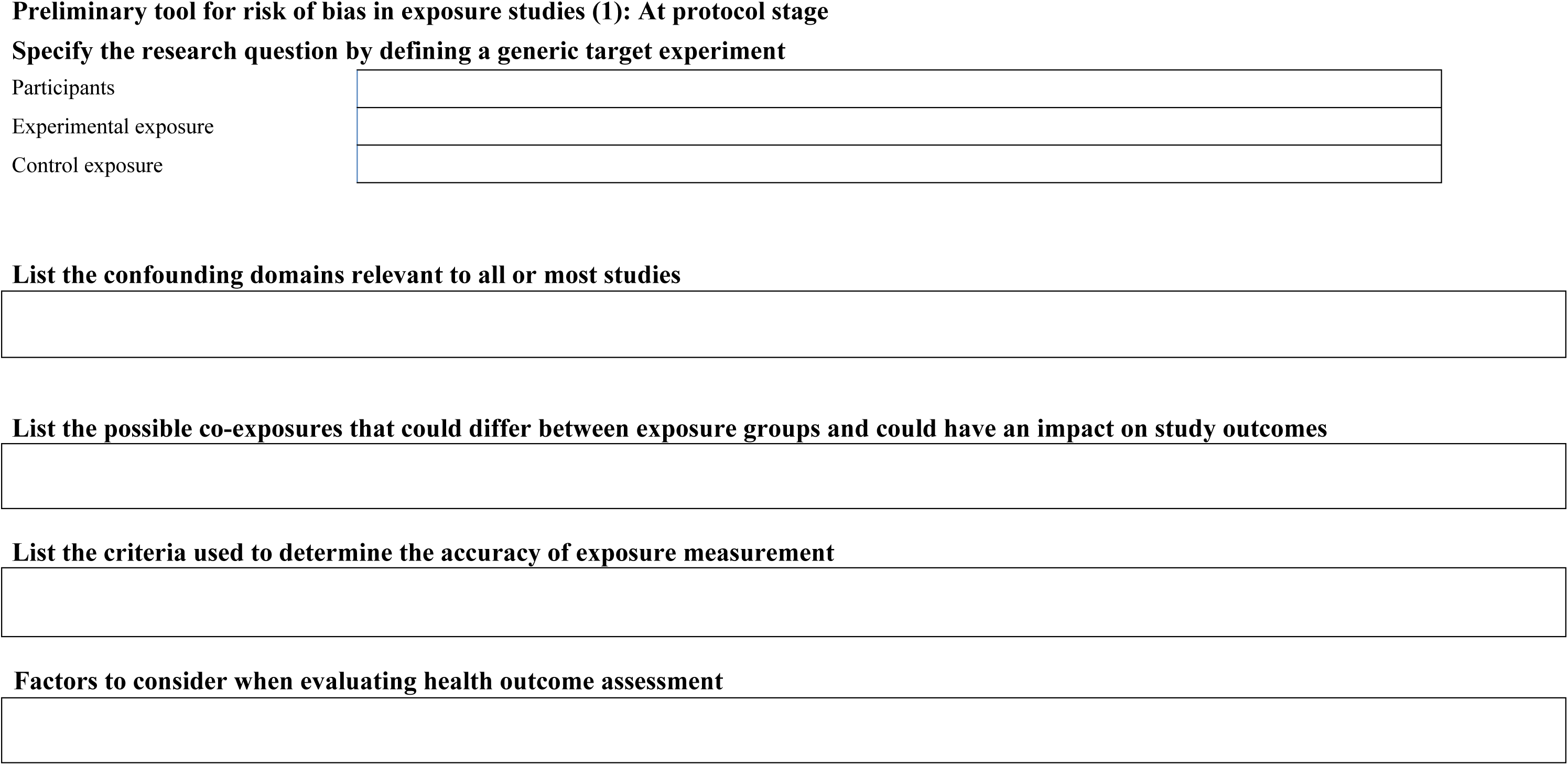
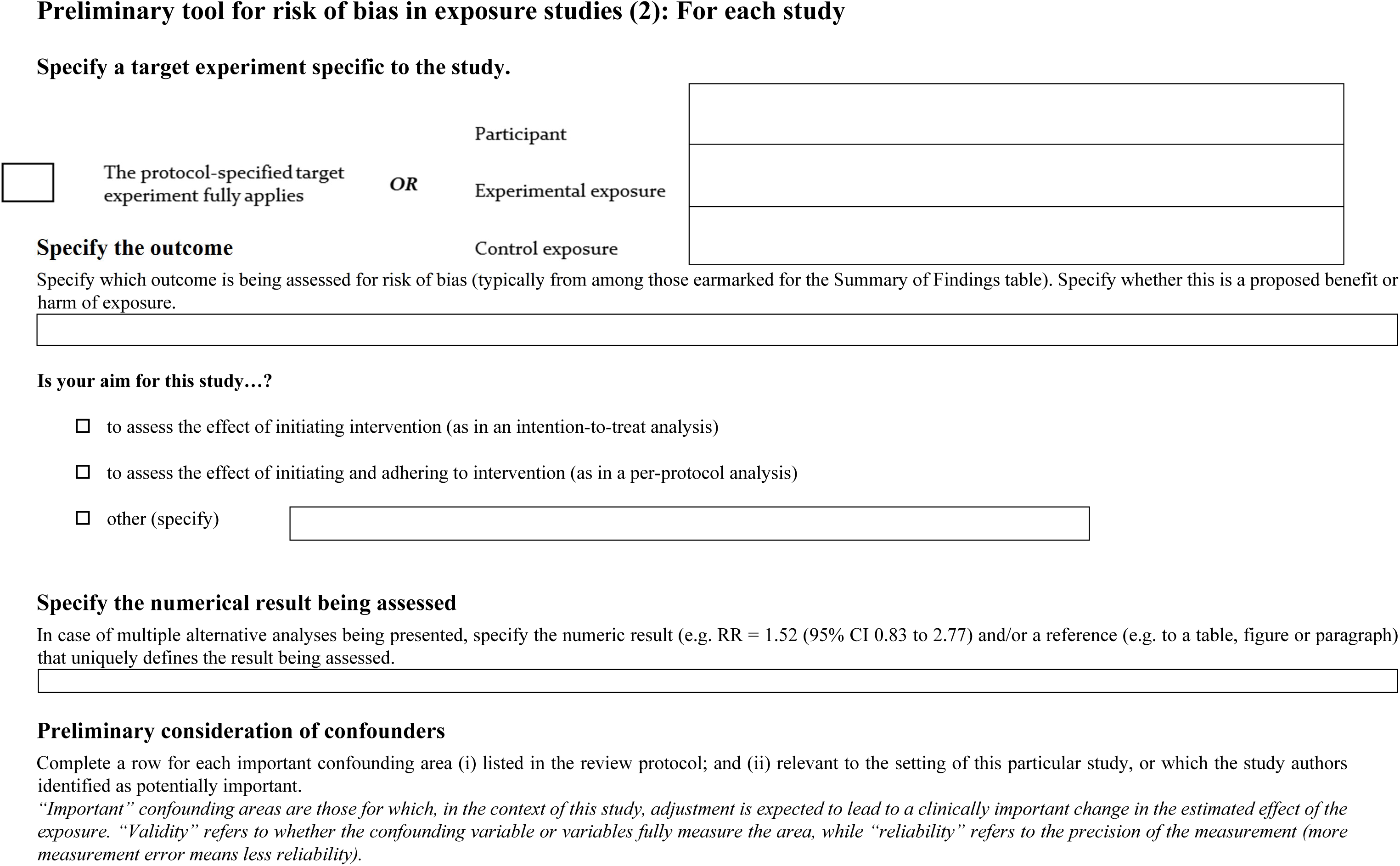
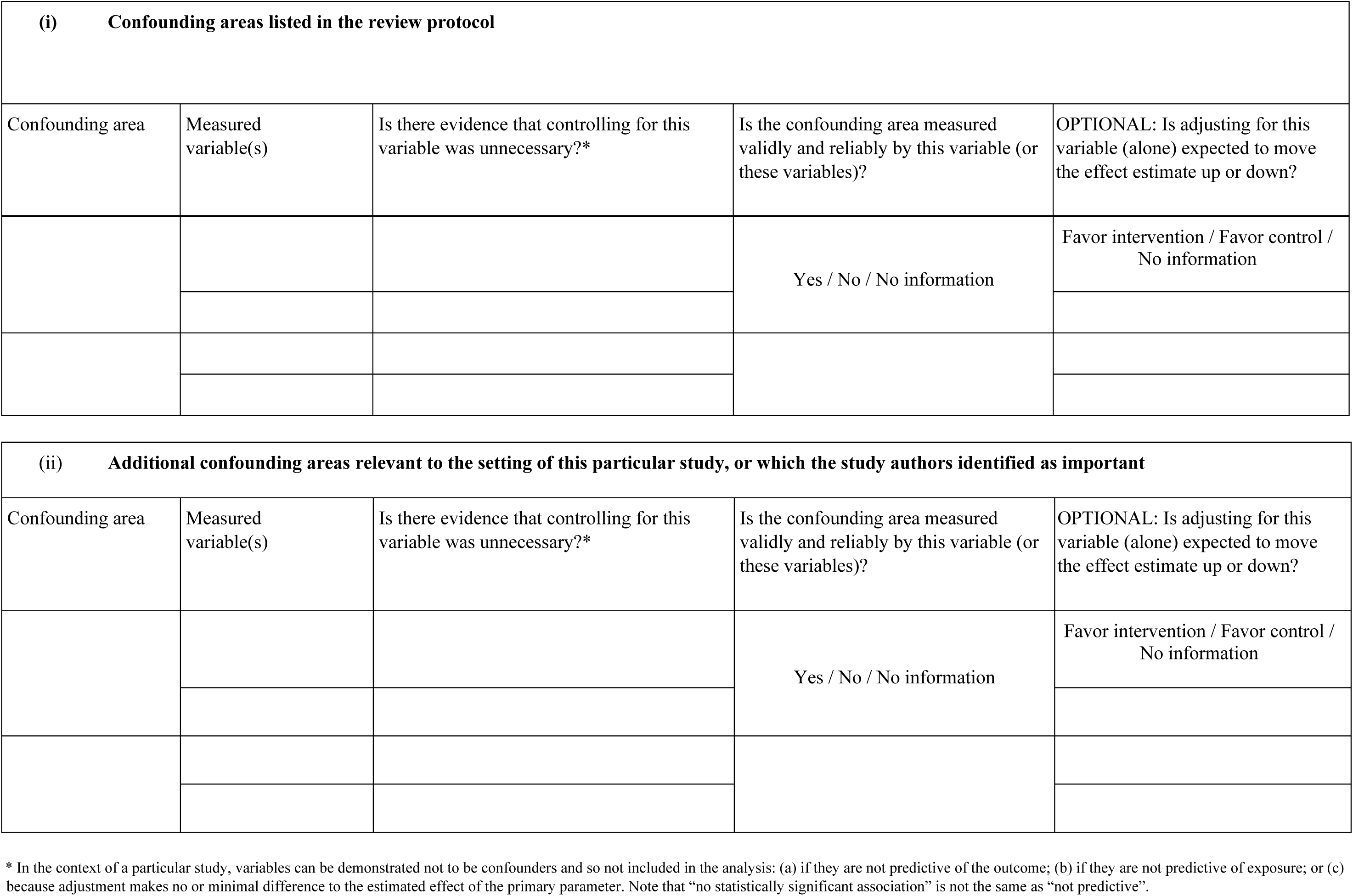
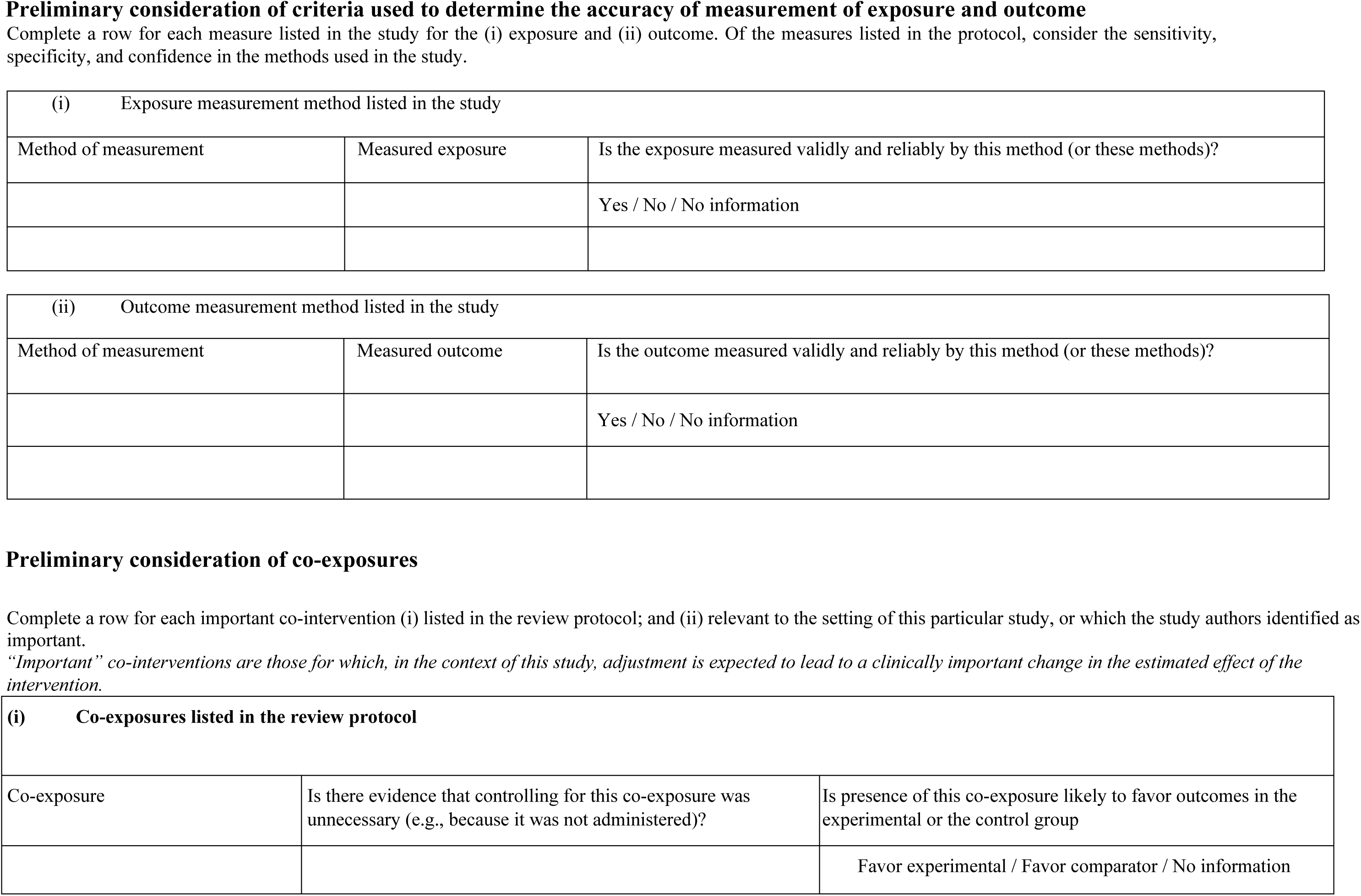
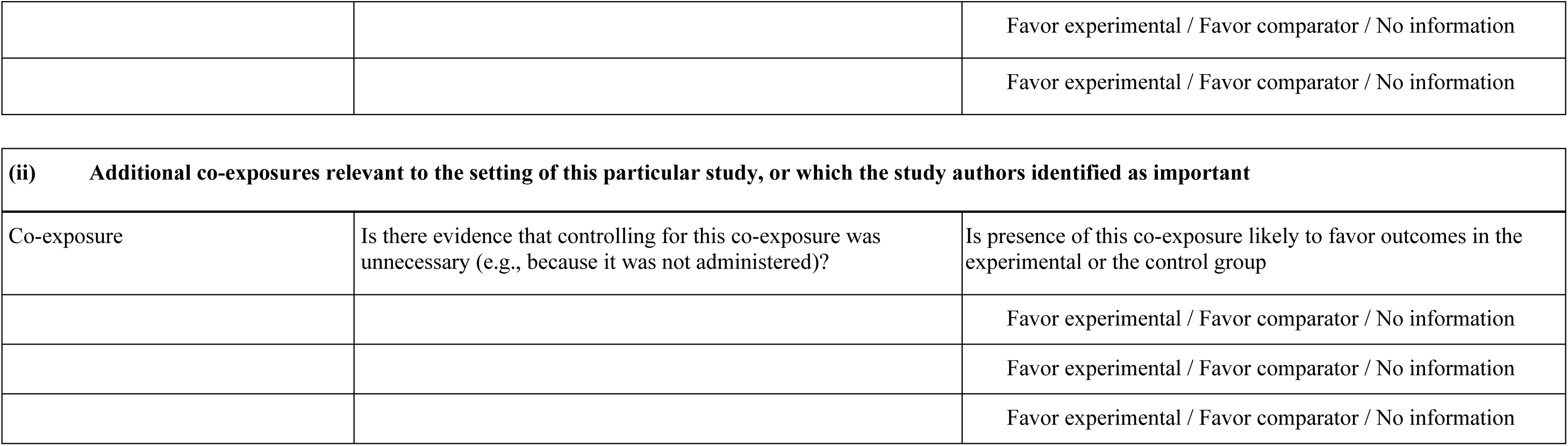
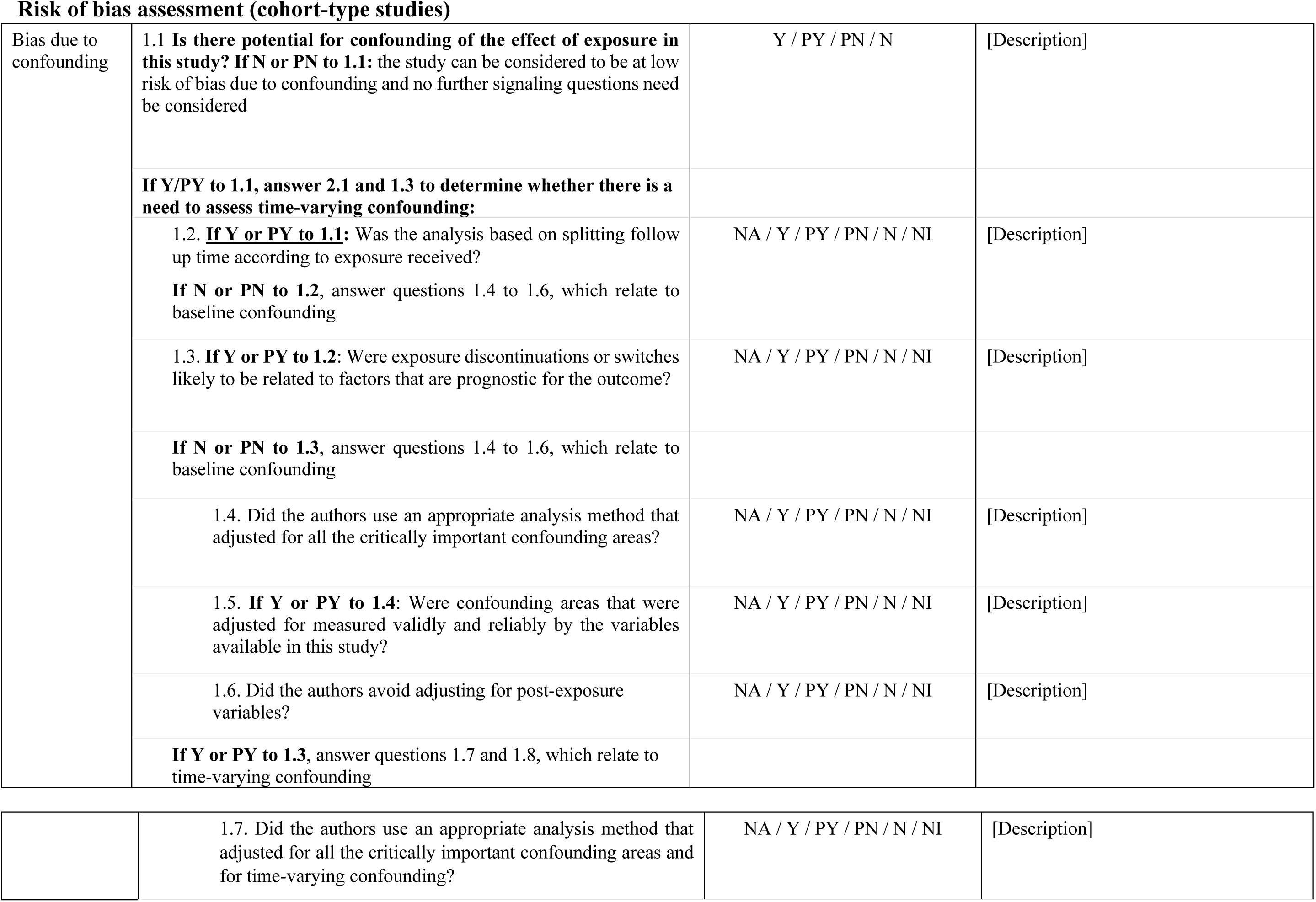
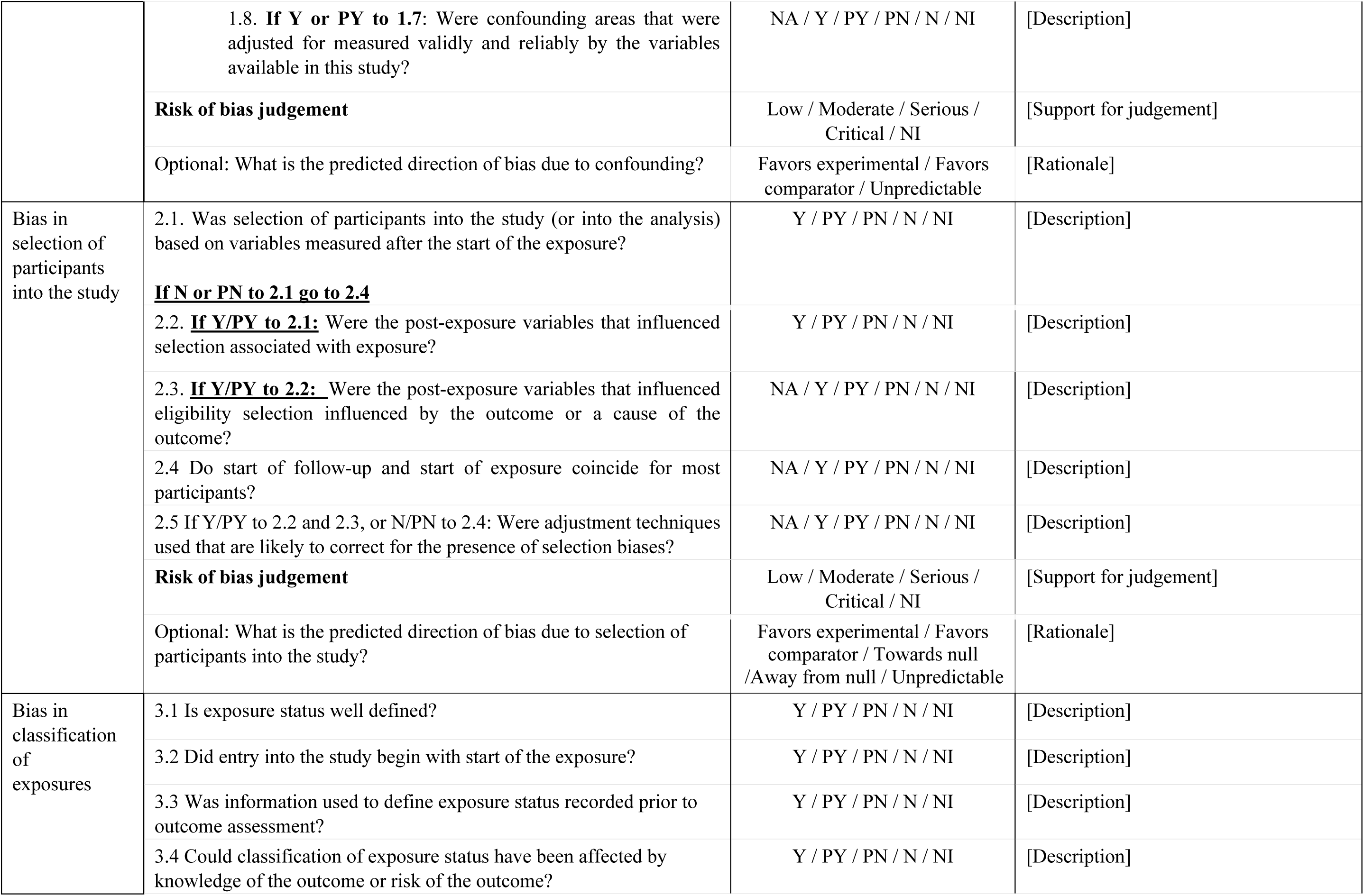
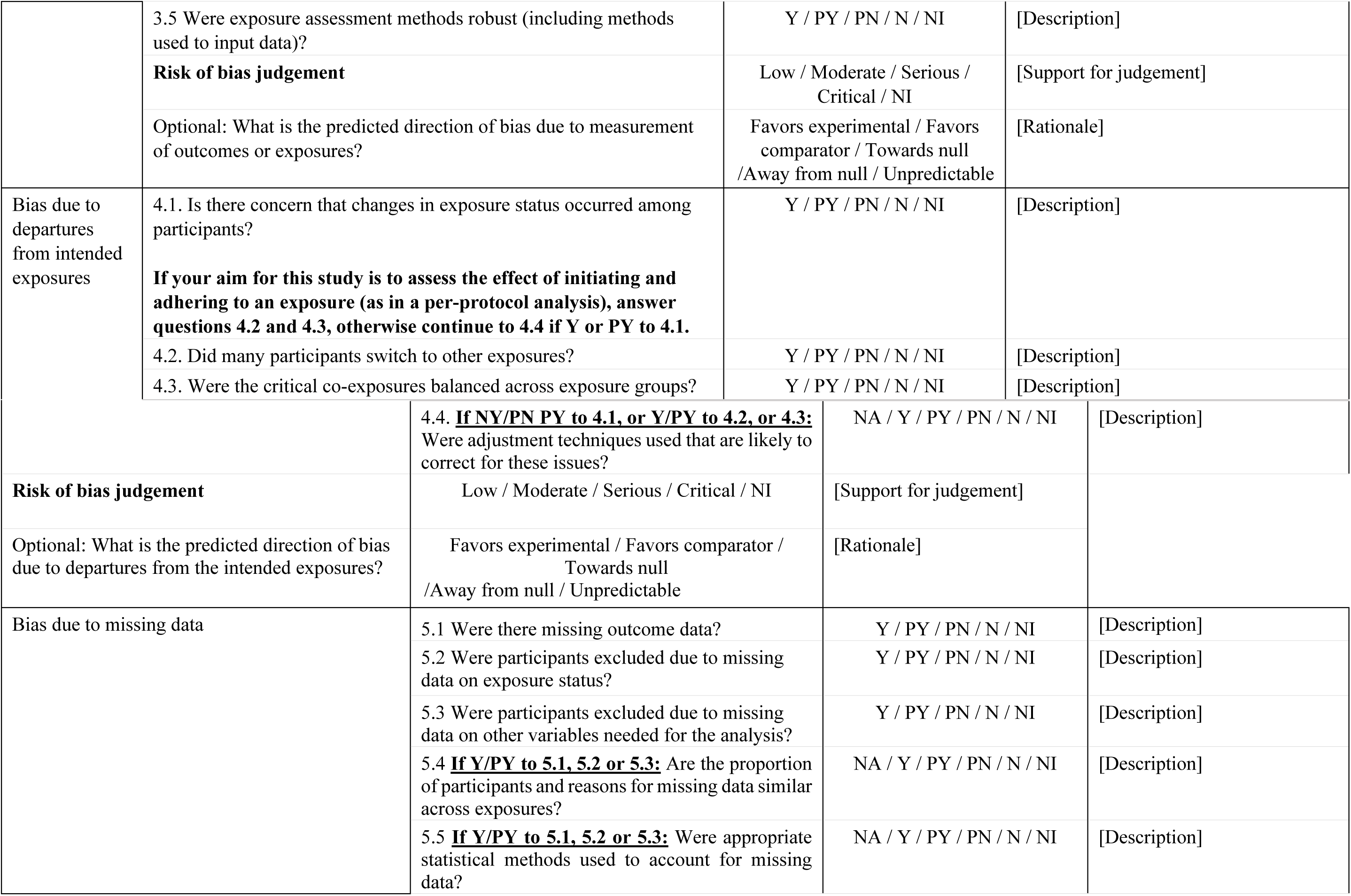
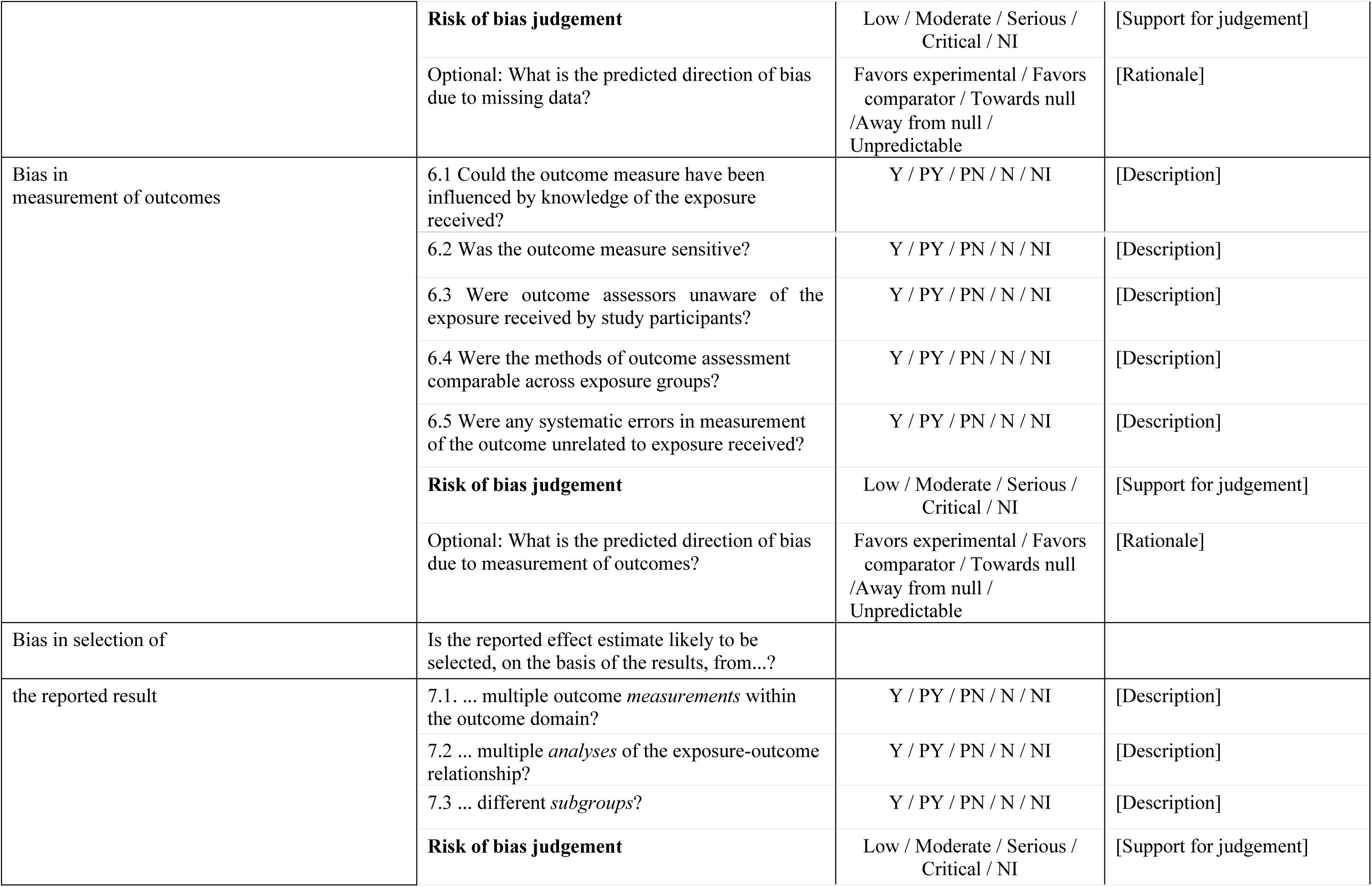
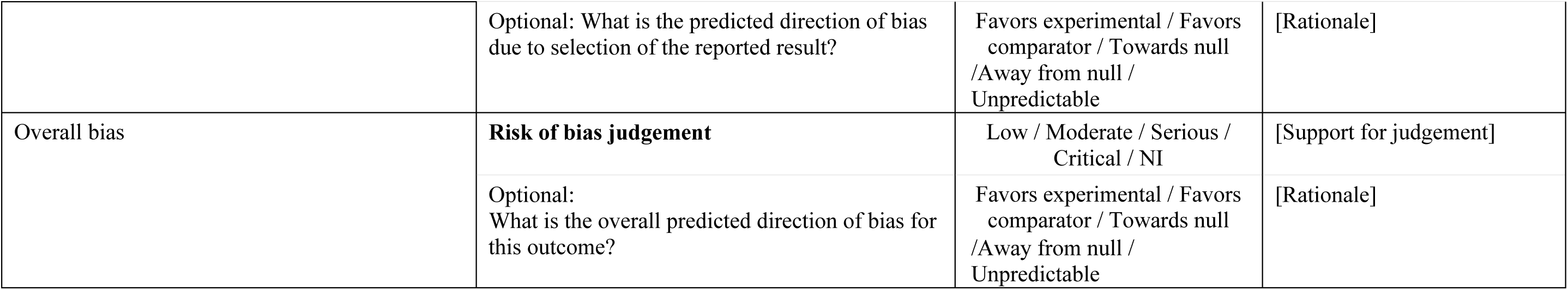

